# Genetic-Driven Rapid and Precise Mimicry of Cardiovascular Fluctuations

**DOI:** 10.1101/2024.07.15.24310460

**Authors:** Minghao Liao, Zhongyou Li, Wentao Jiang, Taoping Bai, Lingjun Liu, Fei Yan

## Abstract

**Background:** Physics-based reduced-order models have gained significant attention for capturing cardiovascular fluctuations. However, achieving quick and precise mimicry of these fluctuations has been a persistent issue for decades.

**Methods:** Inspired by the principle of natural selection, we used a complex whole-body circulation model as an example and utilized genetic algorithms to automate the coordination of model parameters. Additionally, we introduced a “pseudo-distance” metric to evaluate the similarity between the simulated fluctuation curves and the target curves.

**Results:** Through rapid iterations (40 times), this strategy achieved a complete match with the target in both blood pressure and flow fluctuation amplitude and time domains, resulting in highly realistic fluctuation mimicry.

**Conclusion:** This study addresses the major challenge of reduced-order models in the mimicry of blood circulation, ending the history of manual parameter coordination that took months or even years.

**Clinical Perspective:** *What Is New?:* Physics-based reduced-order models are essential for analyzing whole-body hemodynamic status, but they have struggled with complex parameter coordination for decades. This study completely addressed this challenge by employing “genetic algorithms” and an updated “pseudo-distance” criterion, achieving precise mimicry of waveforms both spatially and temporally. Additionally, this work eliminates the dependency on large datasets, making personalized modeling more accessible and practical.

*What Are the Clinical Implications?:* This study lowers the barrier for researchers utilizing these models, significantly advancing the modeling of blood circulation and potentially benefitting physiological analysis, clinical diagnostics, and treatment planning in various cardiovascular events.

## 1.1 Introduction

The circulatory system is fundamental for sustaining human life and activity, as it supplies oxygen and nutrients to the body. Achieving a comprehensive understanding of blood circulation status through systematic, low-cost, precise, and noninvasive methods remains a universally sought-after goal among the global scientific community. Fortunately, the advent of hemodynamic simulations holds transformative significance for gaining insights into hemodynamic phenomena.

With the rapid progress in computing technology, hemodynamic simulation has evolved into a distinct branch of fluid mechanics and is an important part of medical research. Both three- and zero-dimensional simulations are the primary employed strategies ^[1]^. The former is widely employed to capture hemodynamic details, including wall shear stress, oscillatory shear index, and wall deformation of local blood vessels ^[2][3]^. However, mimicking global circulation (whole-body) remains extremely challenging because of the involvement of a myriad of intricate blood vessels, ranging from large to tiny. This process relies on ultra-high-resolution vascular image acquisition and time-consuming three-dimensional reconstruction. Obtaining a well-refined finite-element mesh to ensure computational accuracy is difficult, and even if achieved, the computational demands can be overwhelmingly high ^[4]^. Interestingly, the three-dimensional Navier-Stokes equations can be reduced to a zero-dimensional formulation for pipe flow, analogous to the differential equations for a circuit composed of resistance, inductance, and capacitance. Therefore, a zero-dimensional lumped parameter model (LPM) was developed to mimic cardiovascular fluctuations (blood flow and pressure) through a circuit. Importantly, global blood circulation can be mimicked using closed-loop circuits. This method has been frequently applied to address various cardiovascular issues such as enhanced external counterpulsation ^[2]^, ventricular assist devices ^[5]^, dilated cardiomyopathy ^[6]^, fatigue ^[7]^, hepatectomy ^[8]^, thermal therapy ^[9]^, artificial aortic valve implantation ^[10]^, and hemorrhage ^[11]^. Moreover, the LPM provides more rational boundary conditions for three-dimensional simulations, enhancing the credibility of the computational results. A prominent example is the noninvasive quantification of fractional flow reserve using the 0-3D coupling method proposed by Taylor et al. ^[12]^.

Although the LPM is versatile, its most notable characteristic is the exceedingly high barrier to entry for users due to its multitude of highly coupled and interdependent parameters, making the precise mimicry of cardiovascular fluctuations extraordinarily difficult, as perturbing one parameter can trigger widespread effects throughout the entire circuit. Difficulties associated with the application of the LPM include the need to adjust parameters manually and the continual evolution of the model itself, presenting a long-standing challenge in the field of biomechanics. For example, the C3V-LPM was developed for hemodynamic studies of transcatheter aortic valve replacement ^[13]^, an idealized LPM for Norwood patients ^[14]^, and closed-loop and open-loop models have been proposed to optimize modeling formulas for the left ventricle and systemic circulation ^[15]^. These studies indicated that different models are required for different needs. Still, each instance of model establishment necessitates tedious and inefficient parameter optimization, highlighting the importance of overcoming these issues to improve the applicability of LPMs for specific research purposes. Recently, Li et al. ^[16]^ invested considerable effort into integrating a database of 323 patients by employing a dual neural network to simulate blood flow in the brachial and carotid arteries. However, this approach implies that expanding the simulations to include more localized positions throughout the body requires a significantly larger dataset for effective learning, consequently increasing the workload for data collection and training. This may elevate barriers to entry for users.

Genetic algorithm (GAs) have been incorporated into numerous disciplines, showcasing their ability to efficiently and accurately solve parameter coordination problems. Researchers have utilized GAs to address a variety of challenges, including enhancing the performance of distributed fiber optic sensors ^[17]^, optical wavefront correction ^[18]^, discovering first-order interfacial phase transitions that break the mirror symmetry of asymmetric sigma-tilt grain boundaries ^[19]^, inventing new materials ^[20]^, calculating the optimal magnetization and rigidity pattern that maximizes the workspace in the optimization process of magnetic stimulation soft robots ^[21]^, and designing heat exchangers to minimize the total thermal resistance between the hot fluid and the cold fluid ^[22]^. Furthermore, it is noteworthy that GAs can potentially be perfectly integrated with the parameter coordination of the LPM, as indicated by a few studies in fields such as mechanical and electrical engineering. Examples include the effective structural vulnerability assessment of tubular structures ^[23]^, research on fast self-heating batteries with age-aware consciousness ^[24]^, optimization of the optimal solution for the model of a vertical falling film absorption heat pump ^[25]^, and multi-objective design optimization of current sensor Rogowski coils^[26]^. Drawing inspiration from the successful application of GAs in these critical domains, the integration of this strategy with the persistent bottleneck of parameter coordination in LPMs can be achieved seamlessly.

Unfortunately, there is currently no research integrating GA into LPM. Based on the principles of GAs, we propose that if physiological cardiovascular fluctuations are considered the objective curve, introducing GAs to screen circuit parameters could potentially mimic these fluctuations. This study established a complex model to demonstrate the hypothesis, employing a GA to determine the resistance, inductance, and capacitance (parameters) values of this model. In the GA, we refined the derivative dynamic time warping (DDTW) algorithm to calculate the “pseudo-distance” between the simulated and objective fluctuations, using this calculation as the fitness function to assess their similarity. Ultimately, we established a perfect LPM based on the target waveform information, yielding highly satisfactory bionic results. This study successfully overcame the long-standing challenge of parameter coordination within the LPM, thereby lowering the threshold for users to apply this model to capture global blood circulation, and greatly advancing the cardiovascular modeling. It also provides advanced solutions for designing complex integrated circuits, power networks, and other systems.

## 2 Methodology

### 2.1 Model of global circulation

We constructed a complex LPM for global blood circulation with 131 RLC parameters using the MATLAB platform. This model included the heart, arterial trunk (aorta root, thoracic aorta, abdominal aorta, common iliac artery, external iliac artery, and femoral artery), and branches (celiac trunk, renal artery, mesenteric arteries, lumbar artery, and internal iliac artery) (Fig. 1A). The relationship between ventricular pressure and volume was described using a classic time-varying elastance function ^[27]^. The unidirectional flow through the valves was reflected by diodes. For flow in the arterial trunk and its branches, the associated three-dimensional Navier-Stokes equations can be simplified to one-dimensional equations as follows:

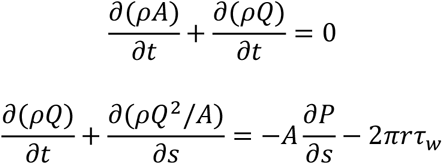

where *ρ, A, Q, P, r, τ*_*w*_ are the density of blood, cross-sectional area of the arterial trunk or its branches, flow rate, pressure, vascular radius, and wall shear stress, respectively. By integrating these equations along the direction of the pipe (*s* direction), the equation can be simplified to a zero-dimensional equation:

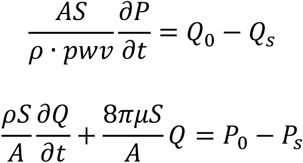

where *S* and *pwv* are the vascular length and pulse-wave velocity, respectively. These two equations represent mass and momentum conservation, and they coincide with the equations formed by electrical resistance (*R* = 8*πμS*/*A*), inductance (*L* = *ρS*/*A*), and capacitance (*C* = *AS*/*ρ* · *pwv*). Therefore, by appropriately determining the *RLC* values, physiological flow and pressure can be obtained.

**Fig 1.**
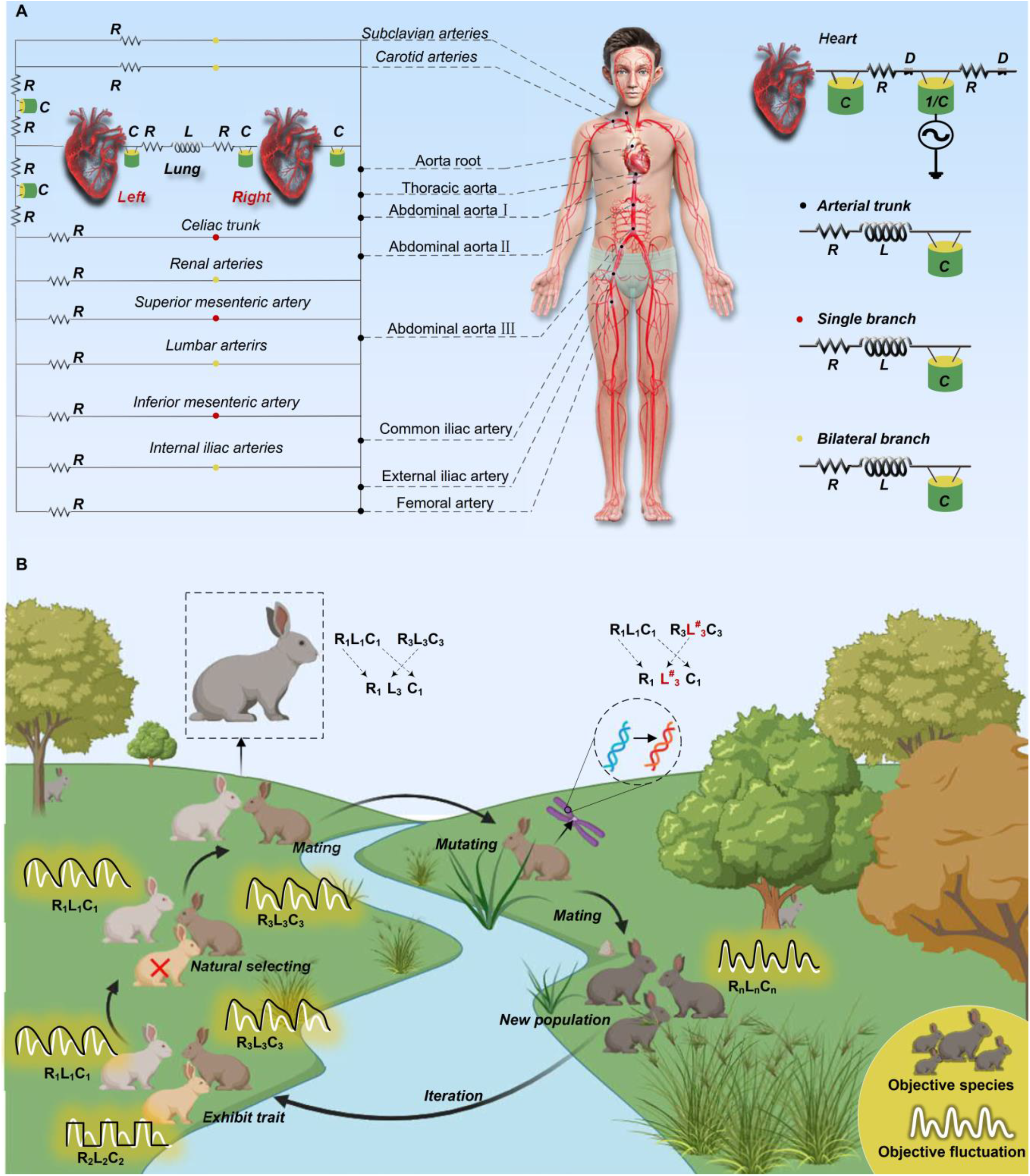
Principle of lumped parameter model (LPM) construction for global blood circulation by genetic algorithm. A, schematic of cardiac and arterial components. Cardiac chambers are modeled by resistor (*R*), capacitor (*C*), inverted capacitor (1/*C*), and diodes (*D*), which represent viscosity resistance caused by the inner wall of the heart, atrial compliance, power source generated by ventricles, and characteristics of unidirectional flow of heart vahje and arterial valve, respectively; the arterial trunk (from the aorta root to femoral artery) and branches (bilateral subclavian, bilateral carotid, celiac, bilateral renal, superior mesenteric, bilateral lumbar, inferior mesenteric, and bilateral internal iliac arteries) are modeled by resistor (*R*), inductor (*L*), and capacitor (*C*), which represent blood flow resistance caused by the arterial wait inertia of blood flow, and compliance of arterial wall, respectively. B, iterative optimization process. Initial parameters (*RLC*) are randomly assigned to each position within LPM, and according to the degree of congruence between their corresponding waveforms and objective waveforms, these parameters are meticulously restructured following the screening process or genetic mutation to formulate new parameter groups. This iterative process continues until modeled waveforms closely approximate target waveforms, which is analogous to the evolutionary adaptation of a rabbit’s coat color to its environment, encompassing the initial exhibition of traits, natural selection, mating, mutation, and generation of a new population.

### 2.2 Genetic algorithm

We used the GA proposed by Holland ^[28]^ to coordinate the *RLC* parameters in this model. The principle of this algorithm is based on Darwin’s natural selection and the genetic mechanisms of biological evolution (Fig. 1B). The GA begins with a new population, and each population comprises individuals with multiple chromosomes. Internally, chromosomes manifest as a combination of genes (genotypes); externally, they manifest as an individual phenotype. Similar to the rabbits in Fig 1B, which continuously evolve and change their skin color to adapt to the environment, the genes carried on their chromosomes (corresponding to *RLC* parameters) determine the skin color of each individual (corresponding to cardiovascular fluctuations). Through the GA, after assigning a series of initial values to the *RLC* parameters, we generated an initial population size of 15 from the search space and followed the principles of survival of the fittest, producing individuals better adapted in each generation. The fitness of each individual’s cardiovascular fluctuations is evaluated using a fitness function and ranked from highest to lowest. Those with the lowest fitness were eliminated, while the fittest fluctuations were selected and combined through genetic operators inspired by natural genetics, such as the crossover and mutation of their *RLC* parameters, to produce a new generation with new *RLC* values. Specifically, the ranges of crossover and mutation rates in our program are 0.3 to 0.9 and 0.01 to 0.05, respectively. If evolution proceeds rapidly after the iterations, the values of the crossover and mutation rates decrease; otherwise, the coefficients increase.

To better integrate the GA into the LPM, we independently develop a MATLAB program, including the main function, subfunction, user-defined model in Simulink, and user guidance (see Supplementary material). This approach offers the advantage of a program interface compatible with Simulink circuit models, eliminating the need to consider the encoding issues typically associated with traditional GA. This allowed us to naturally introduce the parameter values of *RLC* circuit modules as genes into the GA for optimization, leading to higher efficiency. Furthermore, this approach provides greater flexibility for the subsequent optimization of the GA, making it easier for users to customize the algorithm based on their specific needs.

### 2.3 Fitness function

We consider the “pseudo-distance” of fluctuations between the LPM and the objective as the fitness function (similarity) in the GA. Physiological fluctuations differ not only in peak and trough values but also in the timing of these values. As Euclidean distance cannot handle complex time series well, we “warp” the time axis of fluctuations to achieve better alignment by using an updated DDTW strategy. According to the DDTW ^[29]^, consider two time series *Q* and *C*, with lengths n and m, respectively, where:

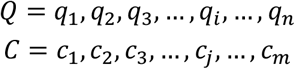

To better “warp” the sequences for comparison, we create an *n* × *m* matrix where the element (*i, j*) records the distance from *q*_*i*_to *c*_*j*_. This element represents the alignment of point *q*_*i*_ with point *c*_*j*_. This n×m matrix is known as the distance matrix *D*, with its (*i, j*) elements denoted as *d*(*q*_*i*_, *c*_*j*_). To avoid the influence of the magnitude of the time series on the DDTW distance, we normalized this distance by:

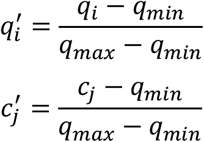

where *q*_*max*_ and *q*_*min*_ are the maximum and minimum values of the *Q* curve. The resulting time series *Q*’ and *C*’ are then subjected to the DDTW algorithm to generate a new distance matrix *D*’ with its (*i, j*) elements denoted as 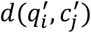.

Based on the distance matrix, we selected a method for aligning the two sequences point-by-point. This method aims to find a path *W* with the shortest cumulative distance *γ*, defined as follows:

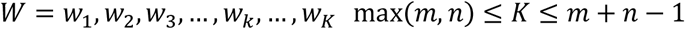

*W* defines the mapping between two time series, where *w*_*k*_ = (*i, j*)_*k*_. The path is subject to the following constraints:

- Boundary Condition: The path must start at *w*_1_ = (1,1) and end at *w*_*K*_ = (*n*, m) within the matrix.
- Continuity: The elements of *w*_*k*−1_ and *w*_*k*_ must be adjacent cells in the distance matrix, including diagonally adjacent cells.
- Monotonicity: Given *w*_*k*−1_ = (*a, b*) and *w*_*k*_ = (*c, d*), it must hold that *a* − *c* ≤ 0, *b* − *d* ≤ 0.

Therefore, for each point (*i, j*), only three directions can be chosen: (*i* + 1, *j*), (*i, j* + 1), (*i* + 1, *j* + 1). The cumulative distance *γ*, with *γ*(*i, j*) representing the sum of the minimum neighboring distances accumulated up to point (*i, j*), is given by:

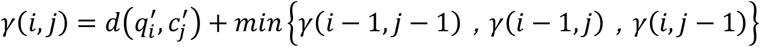

Using this algorithm, we can quickly obtain the shortest cumulative distance, where 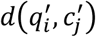 is defined as:

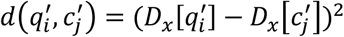

A simple estimate of the derivative was used, which is given by:

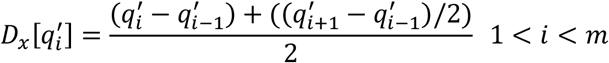

The first and last points were excluded, and the second and second-to-last points were used as proxies for the first and last points, respectively. This estimation is more robust than simply utilizing two data points. However, DDTW focuses more on the similarity of shapes between the time series than on their sizes. Therefore, we redefine 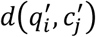 in the distance matrix as the product of the squared difference between the estimated derivatives of points 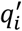 and 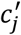, and the squared distance between points 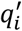 and 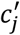, given by:

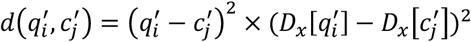

As a result, we use the final cumulative distance *γ* as the distance, or fitness function, between the two time series, and input it into the GA. Evidently, the smaller the value of *γ*, the higher the fitness of the individual, and the more likely it is to be retained for reproduction.

## 1.3 Results

### 1.31 Blood pressure fluctuation

Overall, despite significant initial differences from the target waveform, all pressure waveforms converged toward the target after 40^th^ iteration (Fig. 2). Specifically, the diastolic pressure (<20 mmHg) at the aorta root, thoracic aorta, abdominal aorta I, and abdominal aorta II remained far from the target (Fig. 2A-D), characterized by a large pulse pressure. However, the iterative results were highly satisfactory, with the diastolic pressure adaptively adjusted to nearly normal levels (60-80 mmHg). For cases with small pulse pressures in the abdominal aorta III, common iliac artery, external iliac artery, and femoral artery (Fig. 2E-H), where initial waveforms not only differ greatly in each other (initial values 1 and 2) but also significantly differ from the target, the deviant waveforms nearly overlapped with the target waveform after iteration.

**Fig 2.**
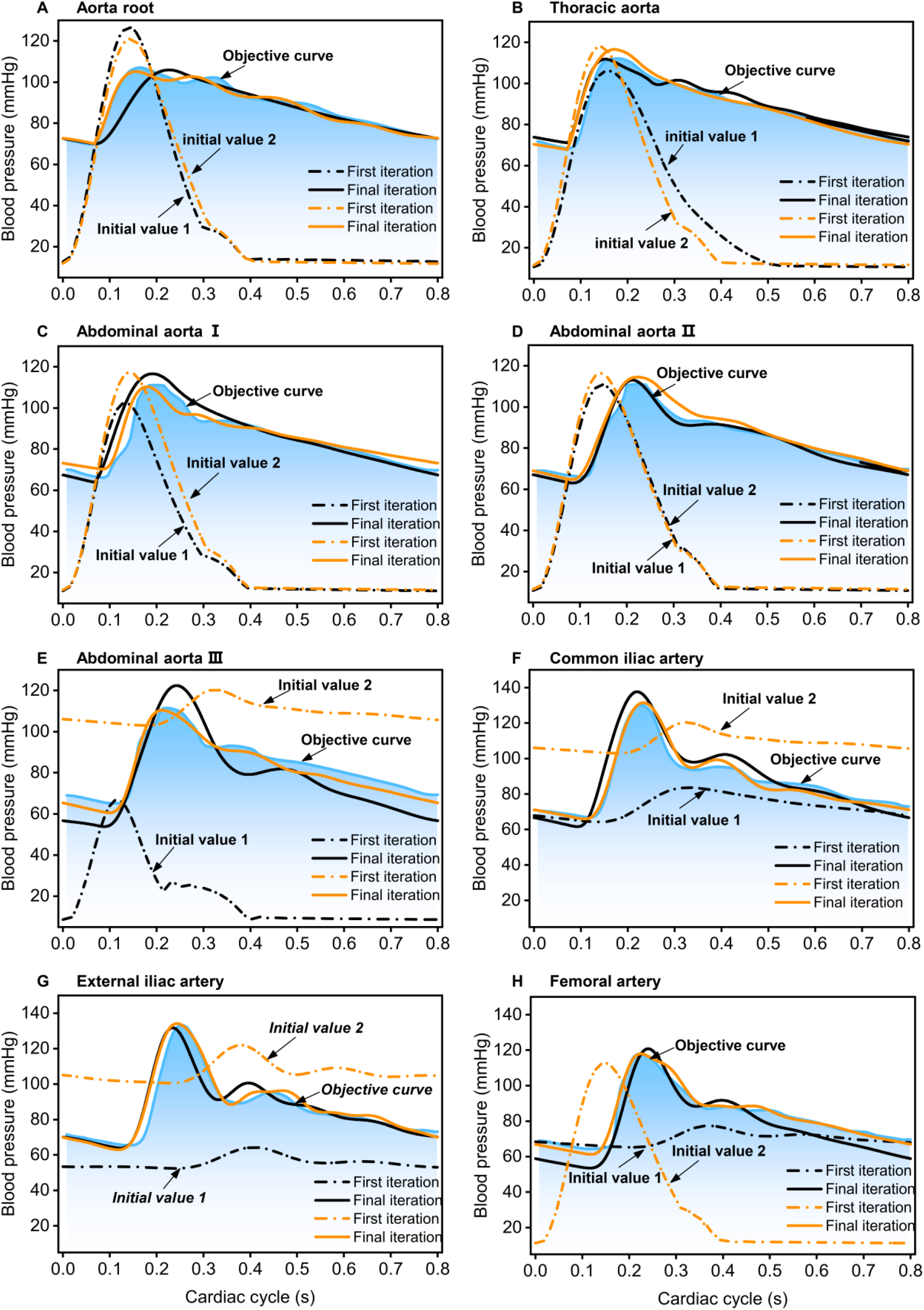
Blood pressure fluctuations of central arteries from aorta root to femoral artery.

### 1.32 Blood flow fluctuation

Both the blood flow waveforms in the arterial trunk and branches converged toward the target after 40^th^ iteration (Figs. 3 and 4). As the key temporal node for flow fluctuation is the amplitude of the peak during the systolic phase, we observed that whether the initial peak exceeded (Figs. 3D, G, H, and 4A, C, D, E) or fell below (Figs. 3B, C, F, and 4B, F) the target, the amplitude converged to a level comparable to that of the target after iterations. It is particularly remarkable that within the same flow fluctuation (Fig. 3A, E), the presence of initial peaks both above and below the target led to similar results. For early diastolic backflow, the amplitude of the backflow after iteration is close to the target, while there is little difference in the flow at the end of diastole before and after iteration. Regardless of whether the initial value is 1 or 2, the proportion of the average perfusion of branch arteries to cardiac output moves from being far from the normal physiological distribution to being close to the target through adjustment via the GA (Fig. 4).

**Fig 3.**
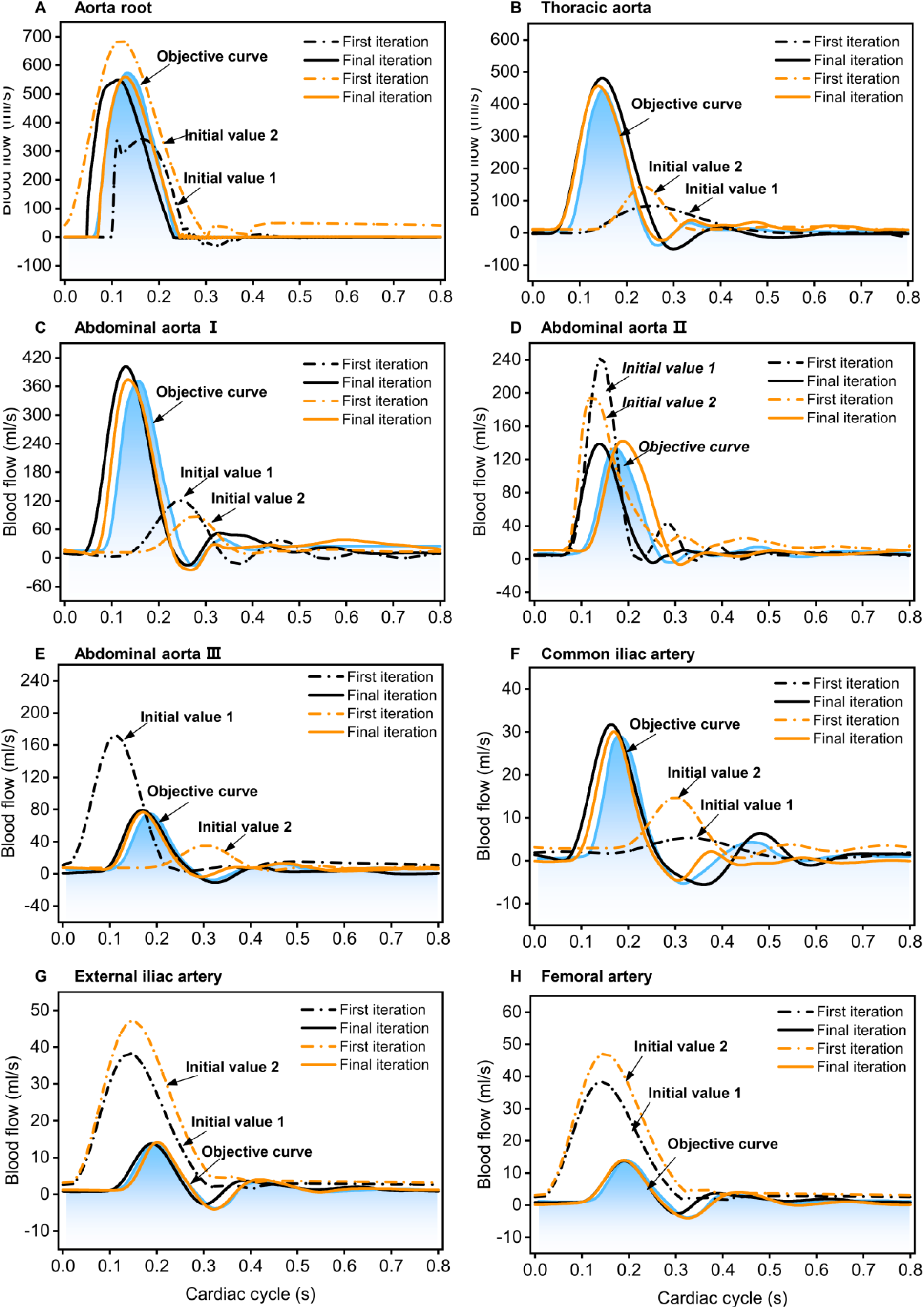
Blood flow fluctuations of central arteries from aorta root to femoral artery.

**Fig 4.**
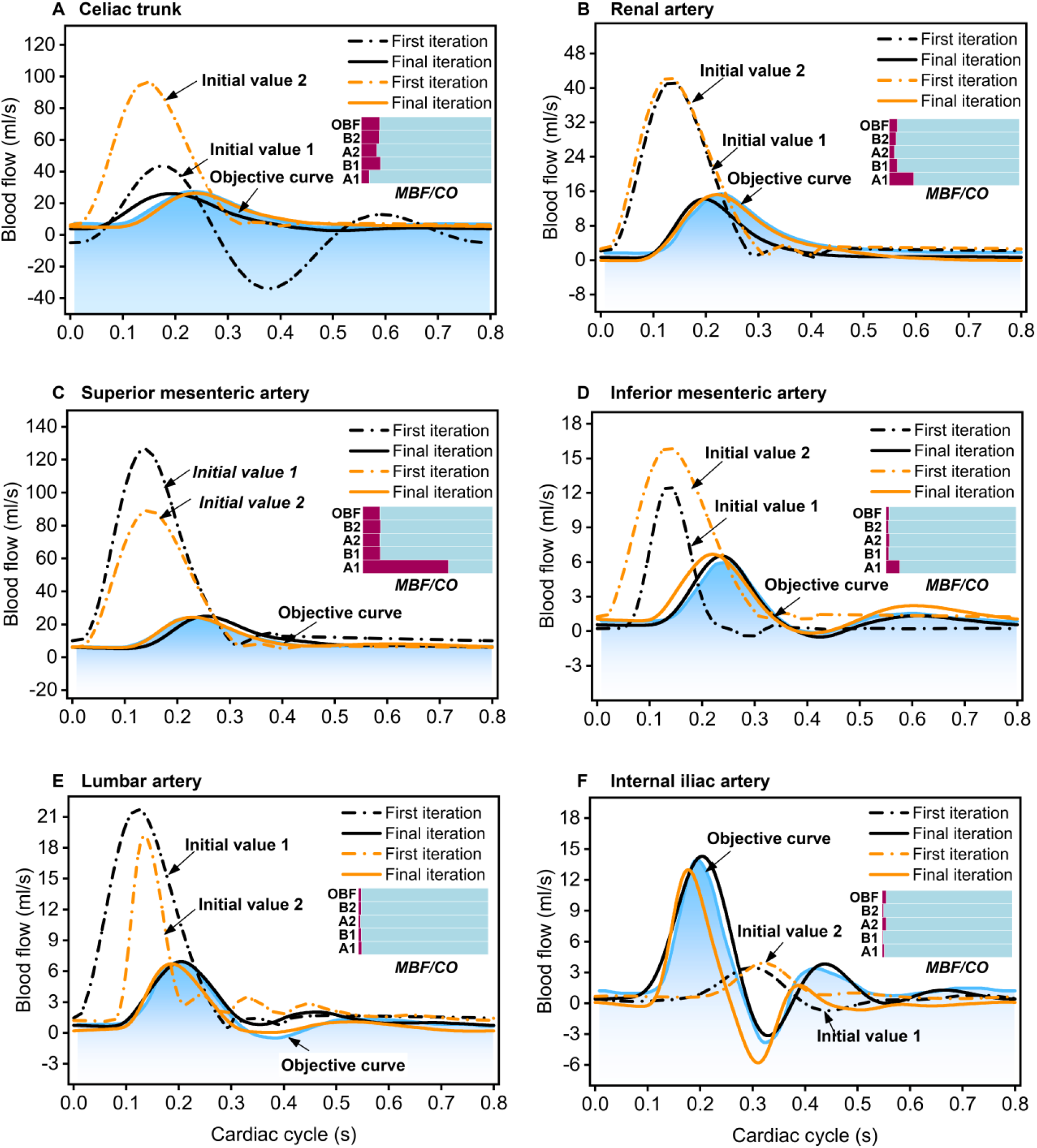
Blood flow fluctuations of branches. The ratio of the mean blood flow (MBF) in each branch to the cardiac output (CO) before and after iterations are also shown in the histogram; A1 and B1, Initial values 1and 2 at first iteration, respectively; A2 and B2, Initial values 1 and 2 at final iteration, respectively; OBF, objective blood flow

### 1.33 Fluctuation patterns of spatial scale

After the final iteration, the systolic pressure from the aortic root to the femoral artery gradually increased (Fig. 5A), aligning with the target pattern, while the diastolic pressure exhibited a decreasing trend (Fig. 5B). This reflects the normal physiological increase in pulse pressure from the heart to the lower legs in the human body. Furthermore, the systolic peak flow from the aortic root to the femoral artery gradually adjusted to an attenuating trend. Between abdominal aortas I and II, where the liver, stomach, and spleen are involved, the attenuation is the fastest, while between the abdominal aortas II and III involving kidneys and intestines, the attenuation is relatively slow (Fig. 5C). The diastolic backflow first increased and then decreased (Fig. 5D), aligning with the target patterns

**Fig 5.**
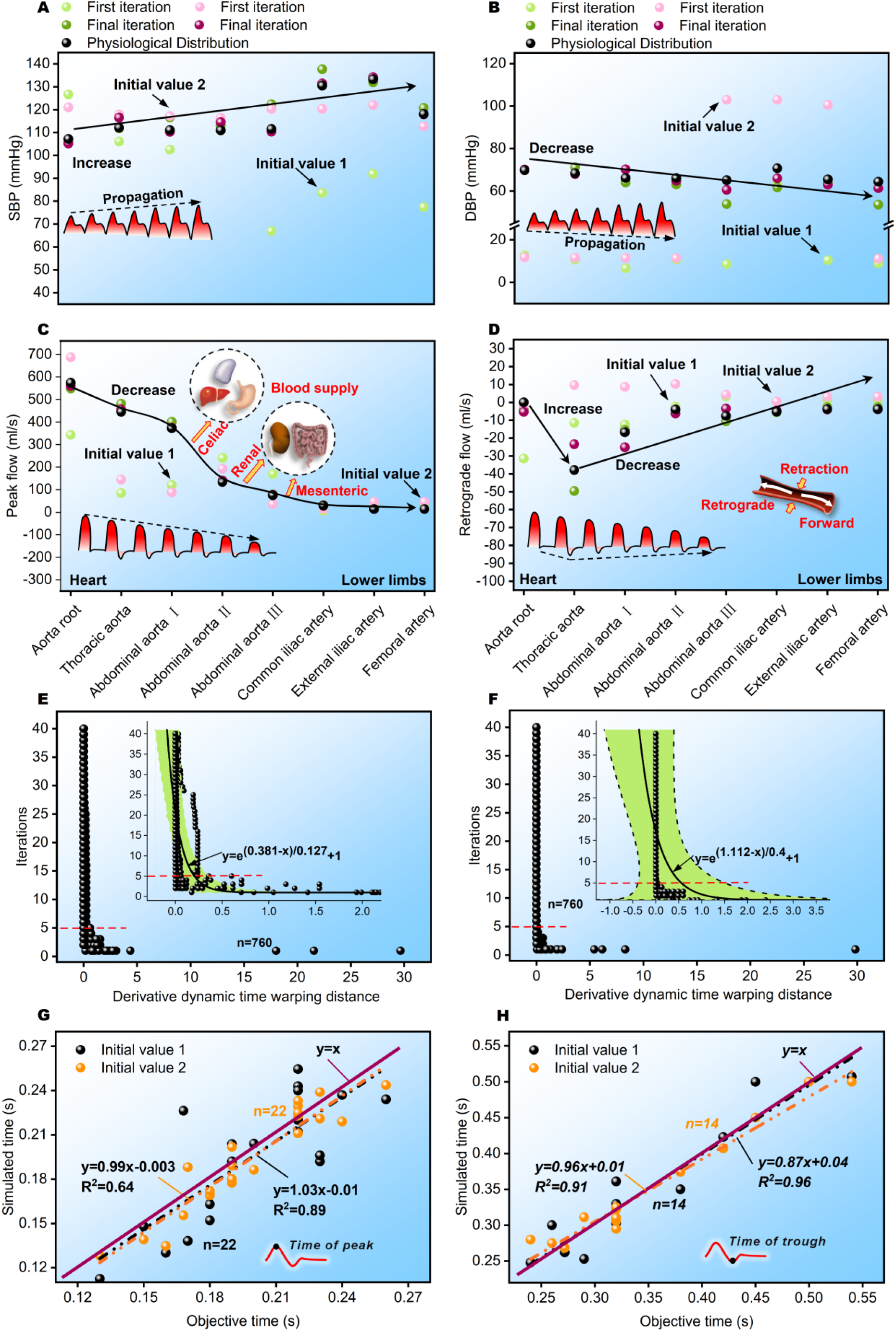
Fluctuation patterns of special moments and similarity of time scale. 258 A and B, patterns of SBP and DBP increase and decreased from aorta root to femoral artery, respectively. C and D, the patterns of systolic peak and diastolic retrograde flow (trough) from the aortic root to the femoral artery, respectively. E and F, the derivative dynamic time warp distance between all simulated and objective curves for initial values 1 and 2, respectively. G and H, the relationship of time points at which peaks and trough appear between simulation and objective, respectively.

### 1.33 Similarity of time scale

For initial conditions 1 and 2, the reciprocal time dynamic twist distance between all simulations and target fluctuations approached zero after five iterations, ensuring similarity in the time series for both sets of curves (Fig. 5E, F). Additionally, under both initial conditions, the convergence speed can be uniformly represented by the function *y* = *e*^(*a*−*x*)/*b*^ − 1 (where *y* is the number of iterations, *x* is the distance, *a* and *b* are constants), indicating that the convergence speed is extremely rapid. Additionally, the special time points between the simulation and the target exhibited a close resemblance. Specifically, the time points at which the peaks (Fig. 5G) and troughs (Fig. 5H) occurred in the simulation showed a linear relationship with those in the target, approximating *y* = *x*.

## 1.4 Discussion

Despite the popularity of reduced-order models based on physics for analyzing blood circulation due to their extremely low amount of calculation ^[6-11]^, the difficulty users face in mastering these methods has been the biggest obstacle to their widespread application. This issue has persisted for several decades.

The core principle of LPM lies in the proper setting of parameters (*RLC*), which is crucial for accurately mimicking physiological fluctuations. However, the current mainstream methods for parameter setting are limited to inefficient manual approaches, which can take months or even years. Even more problematic is the potential need to establish complex models covering numerous regions to meet individualized requirements, involving hundreds of parameters. The coordination of these parameters is extraordinarily challenging because a change in only one parameter can impact fluctuations in many other regions. Therefore, achieving optimal imitation results requires in-depth theoretical study and extensive experience, deterring many users. We attempted to employ artificial intelligence for the continuous learning and output of the desired fluctuations. However, this approach necessitates large datasets for training to achieve accurate results, significantly increasing the workload. Moreover, when considering different research requirements, relearning from large datasets is required, indicating the need for more advanced strategies. For instance, Luca et al. ^[30]^ directly targeted image-based learning and provided an automated method for building reduced-order models. However, they concluded that a sufficiently large training dataset was necessary to achieve a biophysical error in pressure and flow of less than 3%. With this approach, obtaining a comprehensive model of the blood flow of the entire body is practically infeasible because it requires even more complex and high-precision vascular imaging. Similarly, Li et al. ^[16]^ utilized deep learning on the measurement data from 323 individuals (a large sample size) to achieve individualized imitation of brachial and carotid artery blood pressure and flow. Drawing inspiration from the natural selection process, we integrated the GA with an improved curve similarity criterion into LPM modeling to achieve a precise mimicry of complex fluctuations.

The simulation results of blood pressure show that, regardless of whether the initial peak is lower or higher than the target waveform, or whether the longitudinal span of fluctuations (pulse pressure > 100 mmHg) is much greater than the target (30-60 mmHg), all values converge toward the target after 40 iterations, indicating the efficiency of automated modeling. It is known that ensuring overall similarity over the entire timescale is much more challenging than converging to a fixed value. However, using the “pseudo-distance” between the curves as a criterion, we resolved the mutual constraints of the time sequence, allowing the simulated pressure to match the physiological fluctuations throughout the cardiac cycle. Consequently, the variation pattern of the peak pressure from the aorta root (heart) to the femoral artery (lower limb) naturally increases, whereas the diastolic pressure gradually decreases, consistent with physiological conditions [30-33]. This is because there is a decrease in arterial compliance from the large to small arteries, implying self-regulation of the parameter capacitance *C*, which is theoretically reasonable. Moreover, the simulation results for the flow waveforms under different initial conditions were highly satisfactory. Naturally, the ratio of the mean branch flow to the cardiac output approached the target values, indicating reasonable perfusion of the visceral organs. For example, the celiac trunk (11.9% and 13.9% vs. 14.2%, supplying the liver, stomach, and spleen), renal arteries (4.2% and 5.2% vs. 6.2%, supplying the kidneys), and superior mesenteric artery (13.4% and 13.9% vs. 13.3%, supplying the intestines), all exhibited relatively high flow rates because of their critical functions. Additionally, after iterations, the natural decay of peak flow from the aortic root to the femoral artery further indicates the rationality of flow fluctuations, as it involves the blood supply to vital visceral organs, such as the liver, stomach, spleen, kidneys, and intestines. Moreover, the diastolic backflow showed a trend of first increasing and then gradually decreasing from the aorta root to the femoral artery, consistent with previous findings [34]. This is because, near the aorta root, the closure of the aortic valve during diastole prevents blood from flowing backward (acting as a diode). However, due to arterial compliance (capacitance *C*), the arterial wall retraction causes some blood to flow upstream, and this effect gradually diminishes from large to small arteries because of the spatially varying elastic moduli and wall thickness [35].

From the “pseudo-distance” between all curves and the target, the convergence speed exhibits an exponential pattern, indicating an extremely rapid overall convergence. The “pseudo-distance” approaches zero after only five iterations, demonstrating that the two curves become almost identical on the time scale. Consequently, the timings of the peak and trough occurrences are also very similar, which is crucial because they represent the key characteristics of the systolic and diastolic phases, respectively. Therefore, the GA and “pseudo-distance” criterion overcome complex parameter coordination, perfectly fitting the LPM. The simulated fluctuations are precise and rapid, requiring only a given target without the need for extensive clinical samples. Furthermore, based on the success of our model, this method may be equally applicable to adjusting and selecting multi-scale parameters, such as the popular Windkessel boundary conditions ^[36-38]^ and other complex multi-scale coupling studies^[39, 40]^.

However, this study is limited as the simulated curves were not identical to the target curves, which occurred because not all target curves necessarily align with real theoretical solutions. There are always discrepancies between existing measurement data and reality, as well as individual variations. Therefore, the target curves can serve only as references and not as absolute standards.

## 1.5 Conclusion

The strategy of survival of the fittest in GA, combined with “pseudo-distance” as a criterion for curve similarity, effectively addresses the major challenge of rapid and accurate parameter setting in reduced-order models, resulting in the precise mimicry of cardiovascular fluctuations. Notably, this strategy does not require extensive clinical data to establish personalized models, overcoming a long-standing bottleneck in the modeling of blood circulation, and potentially revolutionizing the history of inefficient manual methods. Moreover, this strategy can be beneficial for multi-scale models and various circuit designs.

## Data Availability

The authors confirm that the data supporting the findings of this study are available within the article.

https://github.com/zhongyouli-scu/Genetic-Driven-Rapid-and-Precise-Mimicry-of-Cardiovascular-Fluctuations.git

## Competing interests

The authors declare no competing interest.

## Acknowledgement

This study was supported by the National Natural Science Foundation of China (12302402), the Natural Science Foundation of Sichuan Province (2024NSFSC1377), China Postdoctoral Science Foundation (2024T170616), and the Fundamental Research Funds for the Central Universities (2023SCU12101). We would like to thank Editage (www.editage.cn) for English language editing.

## Author contributions

ZYL, WTJ, TPB, LJL, and FY Contributed to the conception and design of the study; ZYL and MHL were responsible for the acquisition of data, analysis and interpretation of data; ZYL, MHL, and WTJ drafted the article or revising it critically for significant important intellectual content; All authors provided final approval for the version to be submitted.

## Supplementary material

Codes and User Guidance are available at GitHub (https://github.com/zhongyouli-scu/Genetic-Driven-Rapid-and-Precise-Mimicry-of-Cardiovascular-Fluctuations.git).

### Nonstandard Abbreviations and Acronyms

LPM: lumped parameter model
GA: Genetic algorithm
DDTW: Derivative dynamic time warping
MBF: mean blood flow
CO: Cardiac output
OBF: Objective blood flow
SBP: Systolic blood pressure
DBP: Diastolic blood pressure

## Notes

### Competing Interest Statement

The authors have declared no competing interest.

